# Short-stay admissions and lower staffing associated with larger COVID-19 outbreaks in Maryland nursing homes

**DOI:** 10.1101/2021.03.03.21252846

**Authors:** T. Joseph Mattingly, Alison Trinkoff, Alison D Lydecker, Justin J. Kim, Jung Min Yoon, Mary-Claire Roghmann

## Abstract

**Objectives:** Identify facility factors associated with a larger COVID-19 outbreak among residents in Maryland nursing homes (NHs).

**Design:** Observational

**Setting and Participants:** All Maryland NHs.

**Methods:** Resident COVID-19 cases were collected for each Maryland NH from January 1, 2020 through July 1, 2020. Cumulative COVID-19 incidence through July 1, 2020 was collected for each county and Baltimore City. Facility characteristics for each Maryland NH were collected from time periods prior to January 1, 2020. NH outbreaks were defined as larger when total resident COVID-19 cases exceeded 10% of licensed beds. Descriptive and multivariable analyses were conducted to assess the strongest predictors for the primary outcome of larger COVID-19 outbreak.

**Results:** NHs located in counties with high cumulative incidence of COVID-19 were more likely to have larger outbreaks (OR 4.5, 95% CI 2.3-8.7, p<0.01). NHs with at least 100 beds were more likely to have larger outbreaks, especially among facilities with >140 licensed beds (100-140 beds vs <100 beds: OR 1.9, 95% CI 0.9-4.1, p=0.09; >140 beds vs <100 beds: OR 2.9, 95% CI 1.3-6.1, p<0.01). NHs with more short-stay residents (OR 2.2, 95% CI 1.1-4.8, p=0.04) or fewer Certified Nursing Assistant hours daily (OR 2.6, 95% CI 1.3-5.3, p<0.01) also were more likely to have larger outbreaks. Resident race and gender were not significant predictors of larger outbreaks after adjustment for other factors.

**Conclusions:** Large NHs with lower staffing levels and many short-stay residents in counties with high COVID-19 incidence were at increased risk for COVID-19 outbreaks. Understanding the characteristics of nursing homes associated with larger outbreaks can help us prepare for the next pandemic.

**Brief summary:** Maryland nursing homes in counties with a high COVID-19 incidence, more licensed beds, a higher proportion of short-stay residents, or lower CNA staffing hours were more likely to have a larger outbreak early in the pandemic.

## Introduction

Nursing home residents represent a disproportionately high percentage of deaths from coronavirus disease 2019 (COVID-19) for two major reasons. First, they are at high risk for acquiring this infection because of the congregate setting, with proximity to other residents and the staff who care for them. Second, most residents are older adults with multiple comorbidities such as heart disease and diabetes, all associated with higher risks of hospitalization and death from COVID-19.

COVID-19 outbreaks typically occur in nursing homes (NHs) after introduction of the virus from asymptomatic and infectious NH staff, with subsequent spread to other staff, to residents and from resident-to-resident. Infection prevention has focused on a bundled approach for reducing introduction and spread through: visitor restrictions, pre-admission testing and quarantine for residents, routine staff testing, symptomatic resident testing, universal medical masks with eye protection for staff, and cloth face masks for residents when outside of their rooms.^1^ Vaccination of staff and residents will reduce risks even further.^2^

Despite their vulnerabilities, some NHs have had larger outbreaks than others. Studies to date have shown that various organizational and facility factors such as facility size and occupancy, ratings and quality metrics, staffing, and resident ethnicity are related to the presence of COVID-19 in NHs.^3–12^ The purpose of this study was to assess NH factors that determine the trajectory of COVID-19 outbreaks.

We seek to add to the existing literature by focusing on the state of Maryland during its initial wave of infections in the first half of 2020. During this time, the state imposed health directives to restrict visitation, implement infection prevention protocols, expedite testing of symptomatic residents, quarantine new admissions and test all residents and staff in all Maryland nursing homes. Our analysis is unique in that we compare a set of nursing homes under the same health directives by outbreak size and have unique facility data on family satisfaction with nursing home care. This allows us to assess factors associated with outbreak size while controlling for introduction from the local community.

## Methods

### Design overview

This is a secondary data analysis relating facility factors to size of COVID-19 outbreak in Maryland NHs. To evaluate management of the COVID-19 outbreaks, NHs were divided into two comparison groups: those with cumulative resident cases totaling up to 10% of bed capacity, vs those with cumulative resident cases totaling more than 10% of beds from January 1 to July 1, 2020. This was chosen over a binary outbreak definition (yes/no), given that introduction of COVID-19 was likely dependent on community incidence, whereas outbreak size could also reflect facility factors. NHs that contained the outbreak (≤10% of beds) represented facilities that prevented or managed the outbreak effectively.

The sample included all Maryland NHs with Medicare and/or Medicaid Certification that had complete data for all covariates (n=216). Weekly COVID-19 case counts for NH residents were obtained from the Maryland Department of Health coronavirus website^13^ for weeks ending April 29, 2020 (the first week cumulative data from January forward were reported) through July 1, 2020. County-level cumulative incidence of COVID-19 through July 1, 2020 was calculated by subtracting the total number of COVID-19 cases in the county NHs from the total number of COVID-19 cases in the county and then dividing by the total county population. Population and COVID-19 cases by county were obtained from the Johns Hopkins University Coronavirus Resource Center.^14^ Counties were then divided into tertiles by cumulative incidence per thousand population (low: 0-4.00 per 1000, moderate: 4.01-7.86, high: >7.86). Each NH was assigned to the cumulative incidence tertile for its county of location.

NH-level resident and facility characteristics were obtained as a flat file for 2019-20 from the Center for Medicaid and Medicare Services (CMS) CASPER and from the Maryland Health Care Commission (MHCC). These included facility size: small-<100 beds, medium: 101-140, large: >140 beds, ownership: for-profit vs. not for profit (government, other), and CMS quality star ratings, ranging from 1-5. The MHCC defines residents as short stay when facility length of stay is <100 days. Family satisfaction was examined in relation to outbreak size, comparing NHs on the question: “family would recommend facility to others” (<80% would recommend, vs. ≥80% recommend) using the most recent (2017) MHCC family survey data.

Staffing data were calculated as hours per resident day (HPRD) using Payroll Based Journal (PBJ) data from CMS. PBJ data provides daily information on the total working hours of NH staff and the number of residents within a 24-hour period at each NH.^15^ HPRD were calculated by dividing total nursing staff work hours for a 24-hour period by the number of NH residents for the same period during quarter 4, 2019 (October-December).^16^ Then a quarterly average HPRD was calculated for certified nurse assistants (CNAs): nurse aides and certified medication aides. CNA HPRD were dichotomized as <75 percentile vs. >=75 percentile.

### Analysis

Descriptive statistics were reported for all variables, beginning with the unadjusted association between all independent variables and the primary outcome, outbreak size, using student t-test for continuous variables and Chi Square or Fisher’s Exact test for categorical variables, as appropriate. In addition, NHs were compared by COVID-19 outbreak size in relation to the facility factors using multivariable regression techniques. Independent variable selection used forward forced entry, beginning with county-level cumulative incidence, to control for the likelihood of introduction into the facility.^17^ The Hosmer and Lemeshow goodness of fit test^18^ was used to assess model fit. All analyses were performed with Stata 15.1 (Stata Corp., College Station, TX) and SAS 9.4 (SAS Institute, Inc. Cary, NC).

## Results

Of the 227 NHs in Maryland, 216 (95%) NHs were included in our analysis with 6 NHs excluded for missing nursing staffing hours, 4 for missing family experience survey data, and 1 for missing data on age and gender of residents. Table 1 shows the baseline characteristics for residents and facilities as well as the proportion of NHs with total resident COVID-19 cases >10% of licensed beds.

**Table 1.** Nursing home characteristics in Maryland as a baseline in December 2019.

|  | **Maryland**<br>**(N = 216)** | **% or SD** |
| --- | --- | --- |
| ***Resident Characteristics at the Facility-level*** |  |  |
| **Average resident age at facility (Mean/SD)** | 76.2 | 7.8 |
| **Proportion of male residents (Mean/SD)** | 40.1 | 11 |
| **Proportion of black residents (Mean/SD)** | 35.6 | 28.2 |
| **Proportion of residents on Medicaid (Mean/SD)** | 27.8 | 17.5 |
| **Proportion of short stay residents (Mean/SD)** | 61.5 | 18.0 |
| **Proportion of dementia patients (Mean/SD)** | 33.8 | 13.7 |
| **Proportion of patients who are ambulatory (Mean/SD)** | 30.8 | 10.7 |
| ***Facility Characteristics*** |  |  |
| **Total licensed beds** |  |  |
| Small (<100) | 67 | 31.0 |
| Medium (100-140) | 73 | 33.8 |
| Large (≥140) | 76 | 35.2 |
| **Ownership** |  |  |
| Non-profit & government | 53 | 24.5 |
| For-profit | 163 | 75.5 |
| **Chain** |  |  |
| Yes | 116 | 53.7 |
| No | 100 | 46.3 |
| **Alzheimer's unit** |  |  |
| Yes | 71 | 32.9 |
| No | 145 | 67.1 |
| **CMS star ratings – quality** |  |  |
| 1-Star | 0 | 0.0 |
| 2-Star | 17 | 7.9 |
| 3-Star | 39 | 18.2 |
| 4-Star | 77 | 36.0 |
| 5-Star | 81 | 37.9 |
| **Proportion of families who would recommend facility (Mean/SD)** | 79.5 | 15.9 |
| **Certified nursing assistant hours per resident day (HPRD) (Mean/SD)** | 2.1 | 0.4 |
| **Nursing home county-level cumulative incidence of COVID-19** |  |  |
| Low | 28 | 13.0 |
| Moderate | 38 | 17.6 |
| High | 150 | 69.4 |
| ***Outcome Variable: Outbreak size*** |  |  |
| **Total resident COVID-19 cases greater than 10% of licensed beds** |  |  |
| Yes | 127 | 58.8 |
| No | 89 | 41.2 |

Descriptive analyses found associations between several resident and facility characteristics and a larger COVID-19 outbreak (Table 2). Facilities with a higher proportion of residents that were male (p=0.03), black (p=0.0005), and short stay (p=0.004) were associated with having a larger outbreak (>10% of licensed beds). Facility size was associated with COVID-19 outbreak size (p=0.0002) with larger NHs (>140 beds) almost twice as likely to have a larger outbreak as small NHs (<100 beds). Higher CMS quality star ratings were associated (p=0.04) with having a larger outbreak.

**Table 2.** Nursing homes with larger outbreaks: total resident COVID-19 cases greater than 10% of licensed beds through July 1, 2020 as compared to those without.

|  | Total resident COVID-19 cases greater than 10% of licensed Beds |  |  |  |  |
| --- | --- | --- | --- | --- | --- |
|  | No<br>(N = 89) | % or<br>SD | Yes<br>(N = 127) | % or<br>SD | p-value* |
| **Resident Characteristics at Facility level** |  |  |  |  |  |
| Age in Years (Mean/SD) | 77.4 | 8.1 | 75.4 | 7.5 | 0.07 |
| Proportion of residents with dementia (Mean/SD) | 35.5 | 13.4 | 32.5 | 13.9 | 0.12 |
| Proportion of residents who are ambulatory (Mean/SD) | 31.2 | 11.2 | 30.5 | 10.5 | 0.61 |
| **Proportion of male residents** |  |  |  |  |  |
| Less than 40% | 52 | 58.4 | 55 | 44.3 | **0.03\*** |
| Greater than or equal to 40% | 37 | 41.6 | 72 | 56.7 |  |
| **Proportion of Black residents** |  |  |  |  |  |
| Less than 28% | 60 | 67.4 | 55 | 43.3 | **0.0005\*** |
| Greater than or equal to 28% | 29 | 32.6 | 72 | 56.7 |  |
| **Proportion of short-stay residents** |  |  |  |  |  |
| Less than 50% | 27 | 30.3 | 18 | 14.2 | **0.004\*** |
| Greater than or equal to 50% | 62 | 69.7 | 109 | 85.8 |  |
| **Facility Characteristics** |  |  |  |  |  |
| **Total licensed beds** |  |  |  |  |  |
| Small (<100) | 41 | 46.1 | 26 | 20.5 | **0.0002\*** |
| Medium (100-140) | 27 | 30.3 | 46 | 36.2 |  |
| Large (>140) | 21 | 23.6 | 55 | 43.3 |  |
| **Ownership** |  |  |  |  |  |
| Non-profit & government | 26 | 29.2 | 27 | 21.3 | 0.18 |
| For-profit | 63 | 70.8 | 100 | 78.7 |  |
| **Chain** |  |  |  |  |  |
| Yes | 41 | 46.1 | 75 | 59.1 | 0.06 |
| No | 48 | 53.9 | 52 | 40.9 |  |
| **Alzheimer's unit** |  |  |  |  |  |
| Yes | 28 | 31.5 | 43 | 33.9 | 0.71 |
| No | 61 | 68.5 | 84 | 66.1 |  |
| **CMS star ratings – quality** |  |  |  |  |  |
| 1-Star | 0 | 0.0 | 0 | 0.0 | **0.04\*** |
| 2-Star | 11 | 12.5 | 6 | 4.8 |  |
| 3-Star | 16 | 18.2 | 23 | 18.3 |  |
| 4-Star | 36 | 40.9 | 41 | 32.5 |  |
| 5-Star | 25 | 28.4 | 56 | 44.4 |  |
| **Certified nursing assistants HPRD** |  |  |  |  |  |
| HPRD above 75<sup>th</sup> percentile | 32 | 36.0 | 23 | 18.1 | **0.003\*** |
| HPRD below 75<sup>th</sup> percentile | 57 | 64.0 | 104 | 81.9 |  |
| **More than 80% of families would recommend NH** |  |  |  |  |  |
| Yes | 58 | 65.2 | 59 | 46.5 | **0.007\*** |
| No | 31 | 34.8 | 68 | 53.5 |  |
| **Nursing home county-level cumulative incidence of COVID-19** |  |  |  |  |  |
| Low | 22 | 24.7 | 6 | 4.7 | **<0.0001\*** |
| Moderate | 21 | 23.6 | 17 | 13.4 |  |
| High | 46 | 51.7 | 104 | 81.9 |  |

Table 3 shows the results of six different logistic regression models, beginning with our base model that included county incidence, facility size, proportion of short-stay residents, and proportion of families surveyed who would recommend the facility. Across all models, higher county incidence (p<0.0001), larger facilities (p=0.007), higher proportion of short stay residents (p=0.035), and CNA HPRD below 75^th^ percentile (p=0.009) were consistently associated with having a COVID-19 outbreak >10%; race and gender were not significant after adjustment for other factors. A high CMS star quality rating remained associated with a larger outbreak though not significantly so (OR 1.68, 95% CI 0.84-3.35). Family recommendation was somewhat associated with larger outbreaks, though this was attenuated by CNA HPRD. All six models passed the Hosmer and Lemeshow goodness of fit test with p-values above 0.6.

**Table 3:** Logistic regression models to predict Maryland nursing homes with cumulative resident COVID-19 cases totaling >10% of licensed beds, (n=216 facilities).

|  | Model 1 |  | Model 2 |  | Model 3 |  | Model 4 |  | Model 5 |  | Model 6 |  |
| --- | --- | --- | --- | --- | --- | --- | --- | --- | --- | --- | --- | --- |
| Variable | OR | 95% CI | OR | 95% CI | OR | 95% CI | OR | 95% CI | OR | 95% CI | OR | 95% CI |
| County-level Incidence (High vs Moderate/Low) | 3.9 | 2.04-7.48 | 4.38 | 2.24-8.56 | 4.47 | 2.30-8.71 | 4.42 | 2.24-8.70 | 3.67 | 1.76-7.66 | 3.93 | 1.98-7.79 |
| NH size:<br>Medium NH 100-140 bed vs small <100 beds | 2.01 | 0.95-4.26 | 1.84 | 0.86-3.93 | 1.93 | 0.91-4.09 | 1.91 | 0.89-4.08 | 1.8 | 0.84-3.85 | 2.01 | 0.95-4.27 |
| Large NH >140 vs small <100 beds | 2.85 | 1.33-6.08 | 2.72 | 1.26-5.89 | 2.86 | 1.33-6.13 | 2.82 | 1.30-6.12 | 2.63 | 1.21-5.68 | 3.08 | 1.42-6.69 |
| Proportion of short-stay residents (50% or greater vs. <50%) | 2.4 | 1.14-5.04 | 2.27 | 1.07-4.84 | 2.24 | 1.06-4.75 | 2.27 | 1.06-4.86 | 2.50 | 1.14-5.45 | 2.00 | 0.93-4.32 |
| Less than 80% of families would recommend facility | 1.67 | 0.9-3.11 | 1.3 | 0.67-2.54 |  |  |  |  |  |  |  |  |
| Certified nursing assistants HPRD below 75<sup>th</sup> percentile |  |  | 2.33 | 1.10-4.95 | 2.58 | 1.27-5.26 | 2.52 | 1.20-5.32 | 2.49 | 1.22-5.09 | 2.77 | 1.35-5.70 |
| Proportion of male residents below median (40%) |  |  |  |  |  |  | 1.07 | 0.55-2.08 |  |  |  |  |
| Proportion of Black residents below median (28%) |  |  |  |  |  |  |  |  | 1.55 | 0.76-3.15 |  |  |
| CMS star rating: quality = 5 Stars |  |  |  |  |  |  |  |  |  |  | 1.68 | 0.84-3.35 |
| **Bold** if significant at p < 0.05; **Abbreviations:** OR= odds ratio, CI=confidence interval, NH=nursing home, HPRD=hours per resident day, CMS=Center for Medicare and Medicaid Services |  |  |  |  |  |  |  |  |  |  |  |  |

## Discussion

In summary, our analysis found that nursing homes 1) in counties with a high COVID-19 cumulative incidence, 2) with more licensed beds, 3) with a higher proportion of short-stay residents, and 4) lower CNA staffing hours were more likely to have a larger outbreak early in the pandemic. After adjusting for these factors, neither race nor gender were associated with a larger outbreak. The highest CMS star quality rating remained associated with a larger outbreak.

The geographic location of the nursing home had the strongest association with larger outbreaks. This is not surprising, as staff tend to live in the same county where they work and staff introduce COVID-19 into the facility; therefore, adjusting for local prevalence is a proxy for the number of introductions into the nursing home. Community spread is increasingly recognized as a driver for COVID-19 introductions into nursing homes, as county infection rate, population density, and the movement of staff between nursing homes have been described as risk factors for outbreaks.^3,19,20^

Larger nursing homes had a higher risk of larger outbreaks (i.e., cumulative resident cases >10% of licensed beds) perhaps because they employ more staff, thereby increasing the chance of COVID-19 introduction. This is consistent with existing literature suggesting that larger and more fully occupied facilities were more likely to have COVID-19 cases^3–6^. We also found that facilities with a greater proportion of residents who were short-stay had larger outbreaks. Facilities with more short-stay residents have more admissions and, at least early in the pandemic, short-stay admissions from hospitals also may have introduced COVID-19 into nursing homes.

Nursing home metrics reflecting poorer quality, including infection control deficiencies and complaints, have been associated with higher COVID-19 case rates,^3,7,8^ while higher star ratings have been associated with lower rates.^9,10^ Surprisingly, higher CMS Quality ratings were associated with larger outbreaks in our study (see Table 2), perhaps because larger facilities tended to have higher CMS ratings. The highest CMS star quality rating was associated although not significantly with a larger outbreak in our final models.

We did find that facilities with lower family satisfaction scores and fewer CNA staffing hours, two other measures of quality, had larger outbreaks, consistent with existing work.^4,9,11,12^ Higher staffing ratios may lead to better adherence to state-mandated infection guidelines, thus preventing spread, for example, allowing for more rapid adoption of infection prevention practices. Finally, after adjusting for other NH characteristics, gender and race of NH residents was no longer associated with larger COVID-19 outbreaks. This is in contrast to multiple studies suggesting an association between race and COVID-19 cases in NHs, supporting the idea that the NH characteristics (e.g. county location, staffing) are more important than the NHs’ resident demographic characteristics.^3,4,9,10^

Our study is unique as our outcome variable measures the ability to prevent spread of COVID-19 and we account for community prevalence by county in a single state with a coordinated response to COVID-19 in NHs. Most previous studies have used a binary outcome with any case defined as positive,^3–5,7–9,12^ while only a minority have distinguished between small and large outbreaks.^10,11^ We also had a unique measure of quality in the Maryland Health Care Commission’s family satisfaction survey.

In conclusion, our results highlight the importance of local epidemiology in the risk of outbreaks and suggest infection control precautions can be adjusted based on local incidence. Large facilities with a higher proportion of short stay residents are at increased risk for COVID-19 outbreaks despite community infection rates. This highlights the potential importance of separating long- and short-stay residents and the potential advantages of smaller nursing homes for infection prevention through fewer staff members and visitors to introduce infections. Finally, we demonstrate that a high CMS Quality star rating may not be a good measure of a nursing home’s ability to prevent spread in Maryland unlike another quality measure, staffing hours per resident, that was associated with outbreak size. Understanding the characteristics of nursing homes associated with larger outbreaks can help us to better prepare for the next pandemic.

## Data Availability

All data is freely available from the sources mentioned in the manuscript

## Acknowledgements

The sponsor had no role in the design, methods, analysis, or preparation of the manuscript.

